# A population-based serological survey of *Vibrio cholerae* antibody titers in Ouest Department, Haiti in the year prior to the 2022 cholera outbreak

**DOI:** 10.1101/2023.02.06.23285537

**Authors:** Christy H. Clutter, Molly B. Klarman, Youseline Cajusma, Emilie T. Cato, Md. Abu Sayeed, Lindsey Brinkley, Owen Jensen, Chantale Baril, V. Madsen Beau De Rochars, Andrew S. Azman, Maureen T. Long, Derek Cummings, Daniel T. Leung, Eric J. Nelson

## Abstract

After three years with no confirmed cholera cases in Haiti, an outbreak of *Vibrio cholerae* O1 emerged in October 2022. Levels of pre-existing antibodies provide an estimate of prior immunologic exposure, reveal potentially relevant immune responses, and set a baseline for future serosurveillance. We analyzed dried blood spots collected in 2021 from a population-weighted representative cross-sectional serosurvey in two communes in the Ouest Department of Haiti. We found lower levels of circulating IgG and IgA antibodies against *V. cholerae* lipopolysaccharide (LPS, IgG and IgA p<0.0001) in those below 5 years of age compared to those five years and older. Among a subset of patients with higher titers of antibodies, we were unable to detect any functional (vibriocidal) antibodies. In conclusion, the lack of detectable functional antibodies, and age-discordant levels of *V. cholerae* LPS IgG, suggest that populations in Haiti may be highly susceptible to cholera disease, especially among young children.

## INTRODUCTION

In October 2010, the UN peacekeeping mission accidentally introduced cholera into post-earthquake Haiti (1–6). Over the next nine years, approximately 820,000 cholera cases and 10,000 deaths were recorded (7). A response was ultimately organized through a national elimination plan (8), and the last confirmed case of cholera was reported in late January of 2019 (9). In February of 2022, following a 3-year period with no confirmed cases, the Haitian Ministry of Health was optimistic that cholera elimination had been achieved (10). Unfortunately, 10 months later, in a period of instability that compromised access to clean water and sanitation, two cases of cholera were confirmed on October 2^nd^ of 2022 and a new outbreak started, which as of January 31, 2023, had led to more than 27,000 suspected cases and 560 registered deaths (11).

Phylogenetic analyses suggested the current strain was a descendent of the *Vibrio cholerae* O1 Ogawa strain responsible for the original outbreak (12,13). Combatting this new outbreak is challenged by security and humanitarian crises (14,15); however, legacy knowledge and infrastructure remain from the previous outbreak (16,17). There may also be protective immunity afforded by prior infection or vaccination. In the absence of ongoing transmission, younger populations not present in the prior outbreak period would be immunologically naïve and at higher risk of infection. This hypothesis is supported by a report of high rates of young children presenting with cholera (18).

Upon exposure to toxigenic *V. cholerae*, the adaptive antibody response is dominated by two epitopes: cholera toxin and lipopolysaccharide (LPS) (19). However, the most clearly defined non-mechanistic correlate of protection for cholera is the presence of vibriocidal antibodies. These antibodies are functional bactericidal antibodies that target the O-specific antigen of the *V. cholerae* LPS (20–22). Circulating antibody titers peak within several weeks post-infection and slowly wane to baseline over the ensuing months to years, with high levels of inter-individual variability (21). Killed whole-cell oral cholera vaccines (OCVs), such as those distributed in the Haiti vaccine campaigns conducted in Haiti, are on average 58% effective for the first two years, declining to 26% by four years post-vaccination (23). Children under five years of age show approximately half the level of protection by OCV at 2-year follow-up relative to adults (24).

Given that natural infections were not reported, nor vaccinations administered in the three years prior to the 2022 outbreak, we investigated the presence of *V. cholerae-*specific antibodies in adults and children by analyzing samples collected in a cross-sectional serological survey in two communes in the Ouest Department of Haiti conducted prior to the 2022 outbreak.

## METHODS

### Ethics statement

The study was reviewed and approved by the Comité National de Bioéthique (National Bioethics Committee of Haiti; 2021-12), the Institutional Review Boards at University of Florida (IRB202002290), and the University of Utah (IRB_00111137).

### Study design

Cross-sectional serological survey.

### Study population and setting

This study was conducted from February 24^th^ to August 25^th^, 2021, in the communes of Gressier and Leogane, in the Ouest Department of Haiti. This semi-urban and rural region experienced a cumulative attack rate of 5.1-7.5% (25) early in the original cholera outbreak (2010-2012). Cases decreased substantially over time so that towards the end of the outbreak the entirety of the Ouest Department reported 841 cases in 2018 (26). This region was not covered in mass OCV campaigns (27–29).

### Sampling and randomization

Households were randomly sampled geographically and proportionally to population density across the 477 sq km study area. A sampling grid was constructed inside the study area with 149 total grid cells using ArcGIS (ESRI). Grid cells were stratified by population density (low 0-399, medium 400-1511, high 1512-4664) and the sample schema was established *a priori*. Twenty-five grid cells were randomly chosen proportional to population density (2 low, 5 medium, 18 high). An additional 6 low density grid cells were added to have statistical power to conduct future stratified analyses on population density effects. Logistical constraints resulted in 25 grid-cells sampled (6 low, 3 medium, 15 high) out of the 31 intended to be sampled. Within each grid cell sampled, 21 points were randomly distributed and enclosed by Thiessen polygons. Within each polygon, one household was enrolled and sampled; the intention was to enroll from 18 of the 21 polygons per selected grid cell; the remaining 3 were used as ‘back-up’ polygons. The order that grid cells were sampled was randomized.

### Participant recruitment

Haitian staff navigated to each grid cell and polygon (via car, motorcycle, or foot) using ArcGIS Collector (ESRI). The first household reached upon entering the targeted polygon was screened. If the household declined participation or was ineligible, screening continued by proceeding to the next closest house, while remaining within the polygon. A ‘spin the bottle’ app was used to determine the ‘closest’ house if there were multiple equidistant neighboring houses. This process was repeated until either one household was enrolled per polygon (18 total per grid cell) or all households within the polygon were screened.

### Participant inclusion criteria, consent process, and incentives

Eligibility criteria included a household with one adult head of household (HoH) who provided written consent to complete a household survey and at least two household members who provided written consent to provide a dried blood spot (DBS) sample and respond to a household member survey. Parents/guardians 18 years and older provided consent for their minor children and children ≥ 7 years provided assent. Households with a single member were eligible if both consent for the surveys and DBS sample were provided. There was no lower age limit for eligibility. A 500 Gourde ($4.50 US) phone credit was provided to HoH participants to facilitate contact with the study hotline for questions.

### Sample collection

DBS sampling was performed by finger stick, or heel stick in children <1 year, and spotted onto Whatman 903 Protein Saver cards.

### Data collection

The HoH survey queried family demographics and socioeconomic status. The household member survey asked about current and past symptoms and history relating to diarrheal syndromes. Vaccination and disease history were self-reported. Children under 5 years were measured for basic parameters of malnutrition (mid-upper arm circumference (MUAC), weight, and height).

### DBS Eluates

Using a hole puncher, nine 6mm spots were punched from the blood circles on each DBS card and placed into a microcentrifuge tube containing 600 μL of a 1X PBS-Tween solution. The samples were then placed on a tube rocker at speed 50 rpm overnight at 4°C. The next day the samples were centrifuged at 10,500 x g for 2 minutes. The supernatant was removed and stored at -80°C until ready for further use.

### ELISA

Enzyme-linked immunosorbent assays (ELISAs) were performed on all DBS specimens (n=861). Nunc Maxisorp flat-bottom plates (Sigma-Aldrich) were coated overnight with either *V. cholerae* O1 Ogawa-specific LPS (gift of Edward Ryan, Massachusetts General Hospital) or monosialoganglioside (GM_1_, Sigma-Aldrich) at 1 μg/mL. Plates that had been coated with GM_1_ were washed in phosphate buffered saline (PBS) with 0.05% Tween20 (PBST), blocked in PBST + 1% bovine serum albumin (BSA), and coated for a second night in cholera toxin subunit B (CtxB, Sigma-Aldrich) at 2.5 μg/mL. Following coating, plates were blocked, incubated with DBS eluates that were diluted 1:7.5 in PBST + 0.1% BSA, incubated in 1:1000 horseradish peroxidase (HRP) conjugated secondary antibody, either goat-anti-human IgG or IgA (Jackson ImmunoResearch), and developed in TMB substrate solution (Thermo Scientific). Immediately after addition of TMB, plates were placed on a BioTek ELx800 plate reader and read kinetically at 405nm once every minute for eight minutes. Maximum slope (MaxV) values were normalized against a positive control for each plate to generate ELISA units. Positive controls were convalescent plasma from confirmed cholera cases, diluted 1:100. Negative controls were naïve serum (Sigma-Aldrich), also diluted 1:100.

### Vibriocidal antibody assay

The criteria to select samples for vibriocidal antibody assays were those samples with an IgG ELISA titer two standard deviations above the mean for either LPS IgG or CtxB IgG. The assay was conducted via a previously described DBS eluate-adapted drop-plate vibriocidal method (30) and adapted for this study. *V. cholerae* O1 El Tor Ogawa strain (X25049) were grown overnight on thiosulfate-citrate-bile salts-sucrose (TCBS) *Vibrio*-selective agar, after which 2-3 colonies were cultured in Luria-Betani (LB) broth, washed several times with sterile PBS and adjusted to an OD_600_ of 0.3. DBS samples were diluted 1:5 in sterile PBS, heat-inactivated at 56°C, and then serially diluted 1:2 horizontally across a flat-bottom Nunclon™ Delta Surface 96-well plate (Thermo Scientific). Each 96-well-plate included three growth controls and three negative controls. Each batch included at least one monoclonal antibody positive control. Samples and positive controls were cultured for 1hr in a growth solution containing the normalized *V. cholerae* culture (OD_600_ = 0.3) and guinea pig complement (Sigma-Aldrich). Negative controls were cultured with sterile PBS. Plate controls were subsequently plated in 5μL drops onto LB agar. Positive control and participant samples were plated onto TCBS agar (selective media for *V. cholerae*) and cultured overnight at 37°C. Titers were determined as the reciprocal of the first dilution in which the culture drop shows serrated edges or individual colonies.

### Statistical Analyses

Statistical comparisons between two populations were assessed with an unpaired, two-tailed Student’s t test. Significance was determined at p≤0.05. Generalized additive models (GAMs) with a spline to fit age were used to assess the relationship of age and antibody distribution. R version 4.1.3 with the mgcv package was used to fit the splines.

## RESULTS

In this cross-sectional serological survey, we collected DBS samples from 861 enrolled adult (n=564) and child (<18 years, n=297) participants (Table 1); 62.6% were female. A small percentage of participants self-reported prior cholera vaccination (n=10/861, 1.2%) or self-reported clinical disease (n=37/861, 4.3%).

**Table 1.**
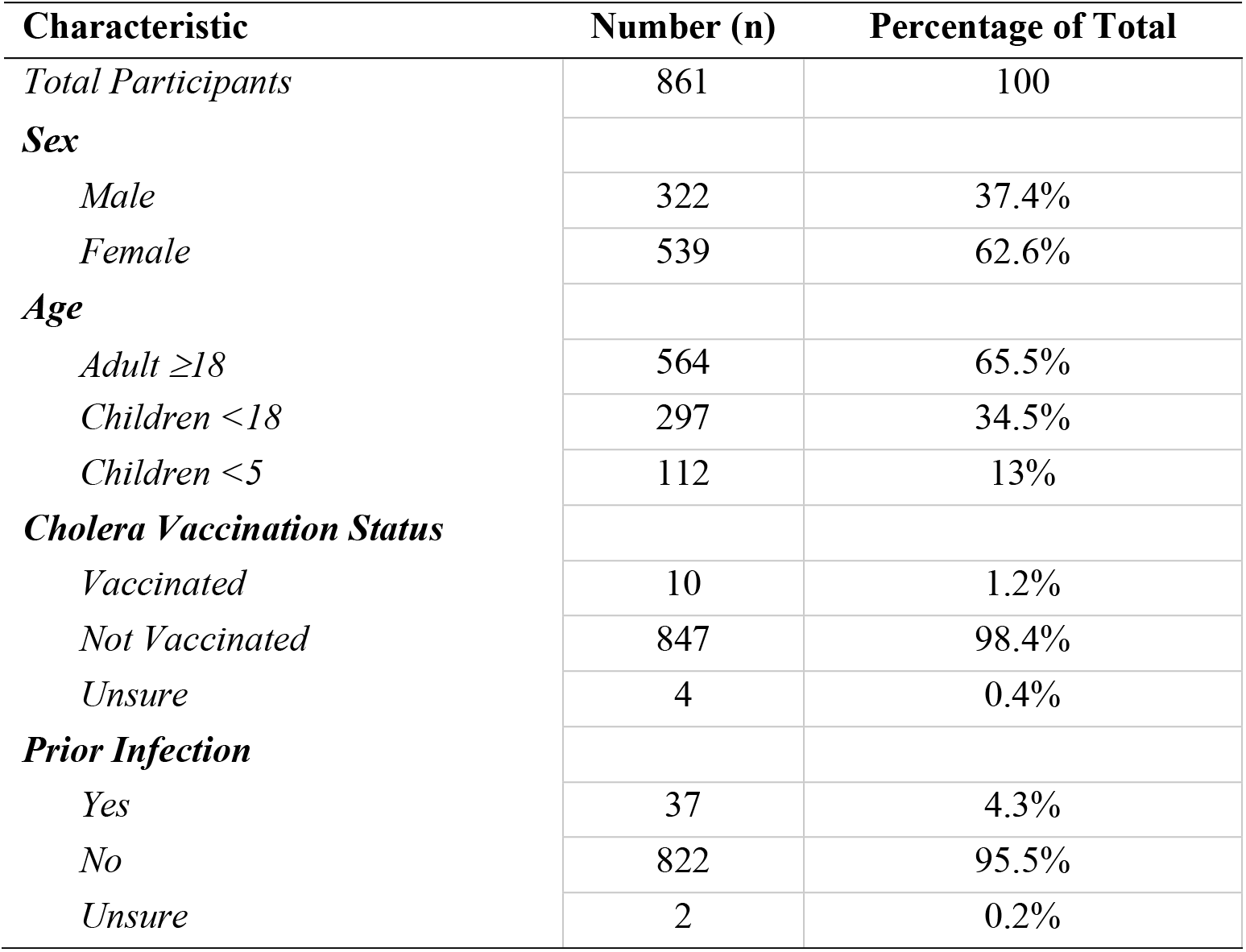
Participant Characteristics.

| **Characteristic** | **Number (n)** | **Percentage of Total** |
| --- | --- | --- |
| *Total Participants* | 861 | 100 |
| **Sex** |  |  |
| *Male* | 322 | 37.4% |
| *Female* | 539 | 62.6% |
| **Age** |  |  |
| *Adult ≥18* | 564 | 65.5% |
| *Children <18* | 297 | 34.5% |
| *Children <5* | 112 | 13% |
| **Cholera Vaccination Status** |  |  |
| *Vaccinated* | 10 | 1.2% |
| *Not Vaccinated* | 847 | 98.4% |
| *Unsure* | 4 | 0.4% |
| **Prior Infection** |  |  |
| *Yes* | 37 | 4.3% |
| *No* | 822 | 95.5% |
| *Unsure* | 2 | 0.2% |

Antibody titers for *V. cholerae* LPS and CtxB were measured across the population for both IgG and IgA isotypes (Figure 1A). Children less than 5 years old had significantly lower titers of both LPS IgG and IgA (p<0.0001, Figure 2A-B, Supplemental Table 1) compared to children and adults 5 and older. CtxB IgG was elevated in children under 5 (p=0.0033), specifically within 1-2-year-old children (p=0.0024 and p=0.0011 respectively, Figure 2C). There was a significant difference between children under 5, older children, and adults with respect to CtxB IgA, but this was driven by the youngest children under 1 year of age, who may lack antibody for reasons unrelated to exposure (Figure 2D). Medians, p-values and interquartile ranges are presented in Supplemental Table 1. Using generalized additive model (GAM) based splines, we estimated a significant positive non-linear association of IgA isotypes with age (LPS: effective degrees of freedom (EDF) 3.4, p<2e-16; CtxB: EDF 2.0, p<e-16) (Supplemental Figure 1). This association was not significant for IgG isotypes (LPS: EDF 4.9, p=0.12; CtxB: EDF 1.0, p=0.13).

**Figure 1.**
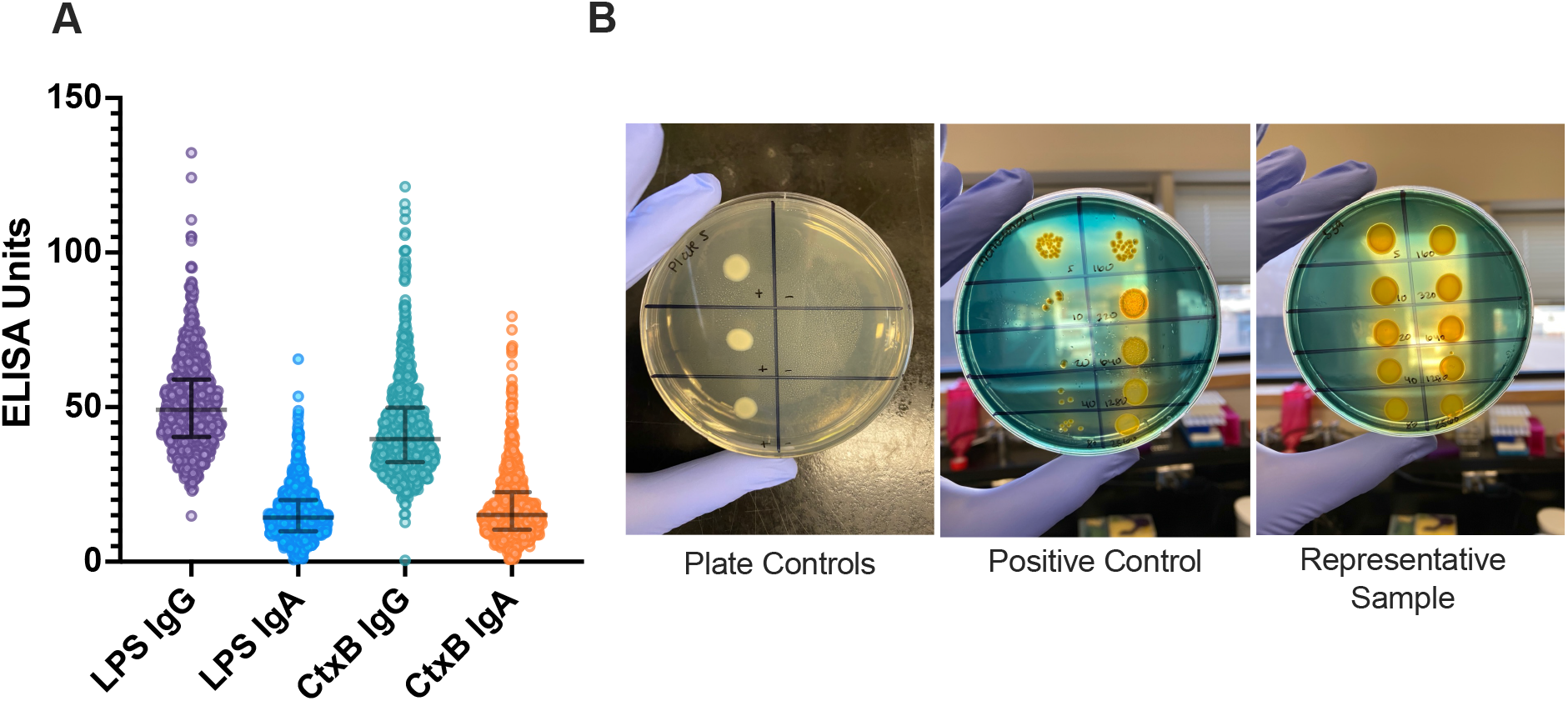
*Vibrio cholerae* specific and functional antibodies. Enzyme-linked immunosorbent assays (ELISAs) were performed on all 861 samples for LPS and CtxB, for both IgG and IgA serotypes (A). Samples that were two standard deviations above the mean for LPS or CtxB IgG (n=51) were assayed for functional (vibriocidal) antibodies (B). Abbreviations: LPS = lipopolysaccharide; CtxB = cholera toxin subunit B. Median and interquartile ranges are displayed (A).

**Figure 2.**
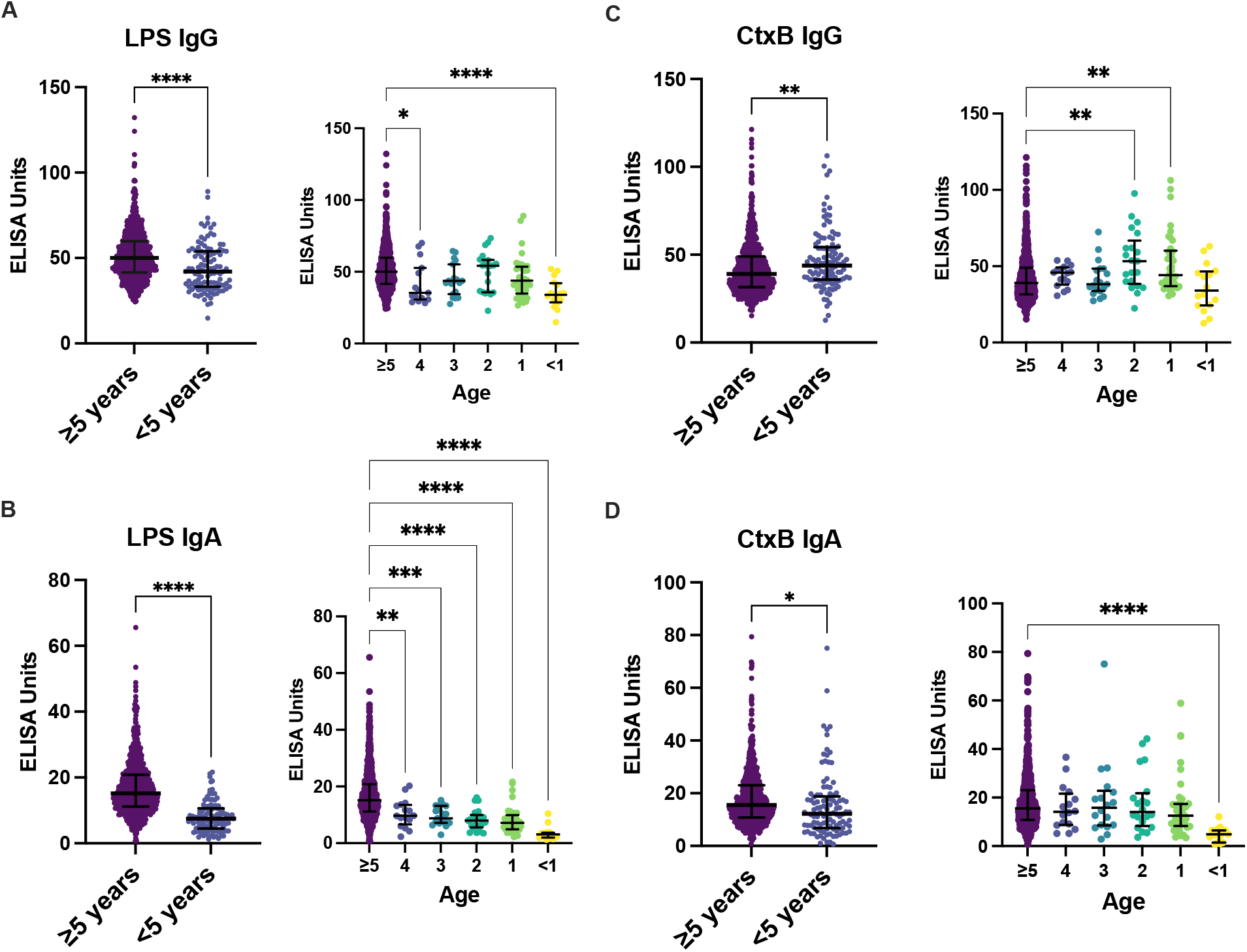
Antibody titers by age, vaccination status, and prior infection. Antibody titers for LPS and CtxB were compared between children under 5 years old (n=112) and adults and children five and older for LPS IgG (A), LPS IgA (B), CtxB IgG (C) and CtxB IgA. Statistical comparisons between ≥5 and <5 were made using an unpaired two-tailed student’s t test. Individual year-by-year comparisons were compared by one-way ANOVA. Significance is annotated as follows: *p≤ 0.05, **p≤ 0.01 ***p≤ 0.001 ****p≤ 0.0001.

Vibriocidal assays were conducted on the subset of participant samples (n=51/861, 5.9%) that had IgG titers for either LPS or CtxB at two standard deviations above the mean. No samples tested had detectable vibriocidal responses (Figure 1B).

## DISCUSSION

In this cross-sectional serological survey in a regional population within Haiti, we detected low rates of circulating IgG and IgA antibodies against lipopolysaccharide (LPS) and cholera toxin subunit B (CtxB). Within the context of increasing antibody response with age, children 1-4 years of age had lower antibody titers against LPS IgG and LPS IgA compared to adults and children over five. Children 1-2 years old had elevated titers of CtxB IgG, which may reflect the cross-reactive nature of CtxB antibodies with the heat-labile toxin (LT) of enterotoxigenic *Escherichia coli*, which has the highest force of infection (measured by seroconversion rate per child-year) among enteric pathogens in Haitian children (31). Due to the inherent cross-reactivity of CtxB, LPS IgG is more specific for exposure history to *V. cholerae* and IgG is more meaningful for age-related comparisons. However, no functional (vibriocidal) antibodies, which is the best available correlate of protection for cholera, were detected.

The serologic assays used in our study have been demonstrated to be associated with prior infection with *V. cholerae* O1 based on longitudinal studies of culture-confirmed cholera patients (21), and also demonstrated to be associated with protection against disease in studies of household contacts of index cases and in human challenge studies (21,32,33). Our data are consistent with limited recent disease transmission and antigenic exposure in this community, especially among young children that were born in the period with little to no circulating pandemic *V. cholerae*. These serologic data suggest that the Haitian communities surveyed may have limited pre-existing immunologic protection against cholera, especially among children under 5 years of age.

The 2022 outbreak was caused by a *V. cholerae* Ogawa isolate that aligns with isolates circulating during the 2010-2019 outbreak (12,13). In the period between outbreaks, the degree to which *V. cholerae* circulated in human and environmental reservoirs at a level below the threshold detectable by the surveillance infrastructure is unknown. The intersection between low levels of circulating cholera and declining population immunity were likely important factors that put Haiti at risk of this new outbreak. These factors, combined with the collapse of clean water and sanitation infrastructure, likely catalyzed the 2022 outbreak.

The findings in this study must be considered within the context of the study limitations. First, there was a risk of enrollment bias because only 28% of the households screened consented to participate, which is lower than other studies in the area that employed similar recruitment methods (34); given oversampling among low density grid cells, the ‘true’ distribution across the population would need adjustment if these data are used as a baseline for future serosurveillance research. Second, the observations of lower IgA antibody titers in children 1-4 may be part of a larger trend of increasing IgA antibody responses with age, representing a confounding factor, and thus may not reflect specific exposures as hypothesized. Third, The ELISA was limited by the availability of quantitative and matrix-matched controls, leading to the use of convalescent plasma as a positive control, and naïve serum as a negative control. Fourth, the selection of samples with high ELISA units for vibriocidal assays may have missed samples with lower antibody levels but harboring functional antibodies. It is possible that matrix differences between DBS and serum/plasma may have impacted the analysis. Finally, our serosurvey was limited to two adjacent communes in the Ouest Department of Haiti, and our findings may not be generalizable to other locations in the country.

## CONCLUSION

In conclusion, our population-based serosurvey of two Haitian communities revealed a lack of functional antibodies, and significantly lower *V. cholerae* LPS-specific IgG antibodies among young children. Our findings suggest that populations in Haiti may be highly susceptible to cholera disease and outbreaks, especially among young children.

## Data Availability

All data produced in the present study are available upon reasonable request to the authors.

## Acknowledgements

We are grateful to the participants as well as the team who conducted the survey in Haiti. We would like to thank the administrators and leadership at the Emerging Pathogens Institute and the Department of Pediatrics at the University of Florida for their ongoing support. We also honor the dedication and support of the Ministry of Public Health and Population (Ministère de la Santé Publique et de la Population - MSPP) for their commitment to the health of the Haitian population despite the profound period of instability.

## Abbreviations and Acronyms

LPS: Lipopolysaccharide
CtxB: Cholera toxin subunit B
DBS: Dried blood spot
ELISA: Enzyme-linked immunosorbent assay
IgG,: IgA Immunoglobulin G, Immunoglobulin A

