## Supplemental Materials for "A population-based serological survey of *Vibrio cholerae* antibody titers in Ouest Department, Haiti in the year prior to the 2022 cholera outbreak"

**Supplemental Table 1. Descriptive Statistics for Age Comparisons**

| Category | Median | Range | 25th Percentile | 75th Percentile | *p* value |
|---|---|---|---|---|---|
| **LPS IgG** | | | | | |
| *≥5 years old* | 50.00 | 107.9 | 41.62 | 59.87 | |
| *<5 years old* | 43.73 | 66.11 | 34.60 | 54.88 | **<0.0001** |
| *4 years old* | 35.36 | 41.87 | 30.66 | 52.66 | **0.0341** |
| *3 years old* | 43.73 | 36.82 | 34.49 | 55.23 | 0.1018 |
| *2 years old* | 54.14 | 50.58 | 35.73 | 58.31 | 0.9633 |
| *1 year old* | 43.77 | 62.41 | 34.80 | 53.59 | 0.0595 |
| *<1 year old* | 33.96 | 37.12 | 28.65 | 42.03 | **<0.0001** |
| **LPS IgA** | | | | | |
| *≥5 years old* | 15.15 | 64.89 | 11.19 | 20.85 | |
| *<5 years old* | 8.075 | 19.43 | 6.038 | 11.20 | **<0.0001** |
| *4 years old* | 9.675 | 17.97 | 6.628 | 13.52 | **0.0052** |
| *3 years old* | 8.770 | 12.18 | 7.140 | 13.16 | **0.0003** |
| *2 years old* | 7.940 | 12.59 | 5.590 | 9.955 | **<0.0001** |
| *1 year old* | 7.175 | 19.43 | 4.930 | 9.928 | **<0.0001** |
| *<1 year old* | 3.075 | 9.100 | 1.953 | 3.730 | **<0.0001** |
| **CtxB IgG** | | | | | |
| *≥5 years old* | 39.12 | 106.0 | 31.55 | 49.02 | |
| *<5 years old* | 45.11 | 83.92 | 36.78 | 55.85 | **0.0033** |
| *4 years old* | 45.77 | 23.11 | 37.98 | 49.23 | 0.9937 |
| *3 years old* | 38.23 | 45.07 | 33.87 | 48.46 | 0.9993 |
| *2 years old* | 53.30 | 75.26 | 38.50 | 66.82 | **0.0024** |
| *1 year old* | 44.22 | 75.73 | 36.96 | 60.15 | **0.0011** |
| *<1 year old* | 34.01 | 50.40 | 24.36 | 46.60 | 0.4507 |
| **CtxB IgA** | | | | | |
| *≥5 years old* | 15.50 | 78.58 | 10.81 | 23.08 | |
| *<5 years old* | 13.95 | 72.12 | 8.620 | 19.77 | **0.0138** |
| *4 years old* | 14.09 | 31.46 | 8.738 | 21.62 | 0.8780 |
| *3 years old* | 15.75 | 72.12 | 8.520 | 22.76 | 0.9979 |
| *2 years old* | 14.03 | 40.64 | 8.278 | 21.80 | 0.9999 |
| *1 year old* | 12.50 | 55.40 | 8.265 | 17.38 | 0.7348 |
| *<1 year old* | 4.900 | 11.41 | 1.440 | 6.470 | **0.0004** |

**Supplemental Table 1. Descriptive Statistics for Age Comparisons.** The median, interquartile range and p-value for each of the age comparisons is shown. Children younger than 5 years were compared in aggregate to children and adults 5 years old and older using an unpaired two-tailed student t test. Individual age groups of 1, 2, 3, and 4-years were compared to older children and adults by one-way ANOVA. Units for all columns excluding the p-value refer to ELISA units.

**Supplemental Figure 1. ELISA Units analyzed by age of participant.**

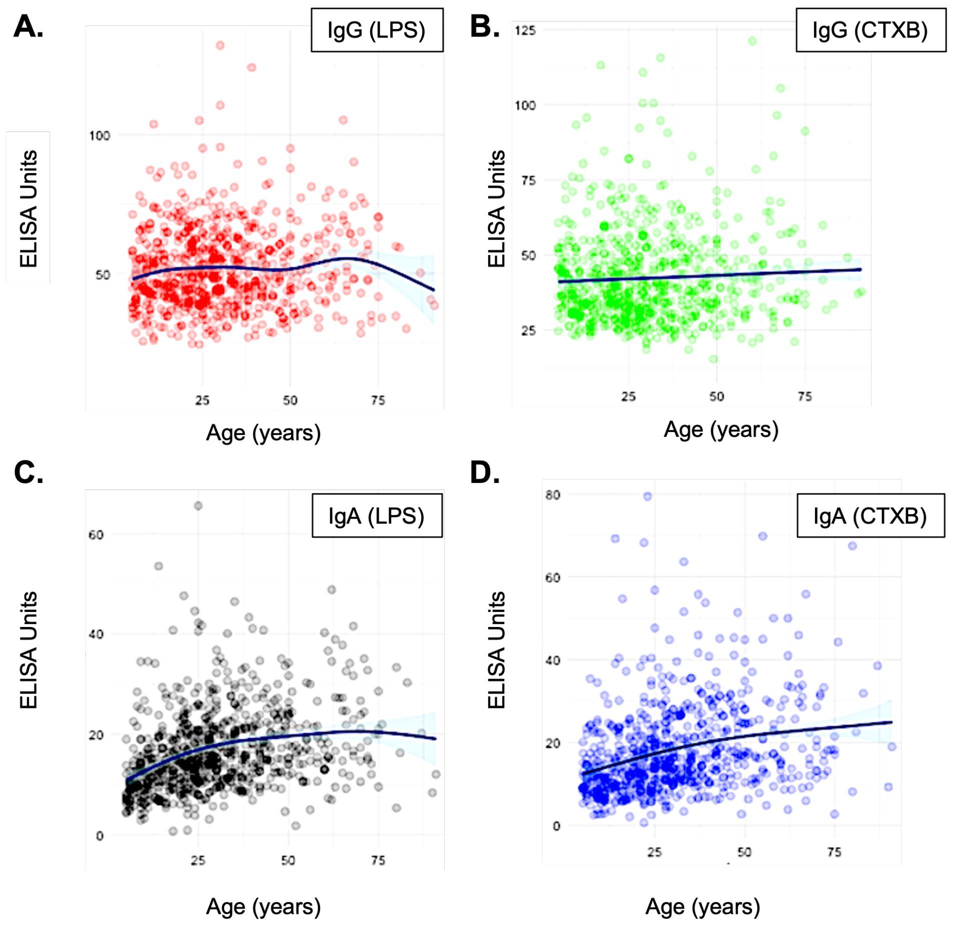


**Supplemental Figure 1.** Antibody levels expressed in ELISA units analyzed by age of participant using a generalized additive model (GAM). **A.** IgG to *V. cholerae* LPS (effective degrees of freedom, EDF, 4.9, p=0.12); **B.** IgG to *V. cholerae* CtxB (EDF 1.0, p=0.13); **C.** IgA to *V. cholerae* LPS (EDF, 3.4, p<2e-16); **D.** IgA to *V. cholerae* CtxB (EDF 2.0, p<2e-16)**.** A statistically significant association (p<0.05) was identified between age and IgA for both LPS and CtxB.
